# Clinical performance of two EUA-approved anti-COVID-19 IgG/IgM rapid lateral flow immunoassays using whole blood finger-sticks

**DOI:** 10.1101/2021.06.04.21258189

**Authors:** Christian Tagwerker, Irfan Baig, Eric J. Brunson, Kristine Mundo, Davan Dutra-Smith, Mary Jane Carias-Marines, Ranulu Samanthi de Zoysa, David J. Smith

## Abstract

Serological, or antibody, tests detect immunoglobulins produced by the hosts plasma B cells following exposure to foreign antigens. Venipuncture blood draws to collect human venous whole blood, plasma from anticoagulated blood (Li+ heparin, K2EDTA and sodium citrate), or serum are commonly utilized and require refrigerated temperatures during transport to the testing facility. Subsequent laboratory testing by enzyme-linked immunosorbent assays (ELISA) or chemiluminescence immunoassays (CLIA) can take an additional 2-5 hours. In the context of the COVID-19 pandemic, rapid diagnostic tests (RDT) to be used in point-of-care (POC) and remote settings have become essential during mandatory quarantine and isolation periods. RDTs allowed for more cost-effective testing using less collection materials with an immediate (5-10 minutes) test result. However, the majority of emerging RDTs receiving Emergency Use Authorization (EUA) approval by the Food and Drug Administration (FDA) for qualitative detection and differentiation of IgM and IgG antibodies to SARS-CoV-2 were only approved for use in human venous whole blood, plasma or serum. In this study we summarize performance characteristics of one RDT (COVID-19 IgG/IgM lateral flow immunoassay rapid cassette) to another by simultaneous application of whole blood finger-stick specimens (n = 32). The study was performed over 5 different days, with daily quality controls consisting of serum previously verified to be positive or negative by COVID-19 IgG/IgM ELISA testing.

## 1 Introduction

An outbreak of respiratory disease caused by a novel coronavirus SARS-CoV-2 (COVID-19) first detected in Wuhan City, Hubei Province, China, has demonstrated the capability to spread rapidly, leading to significant impacts on healthcare systems and causing societal disruption was designated as a pandemic by the World Health Organization (WHO) [1]. To respond effectively to the COVID-19 outbreak, rapid detection of cases and contacts, appropriate clinical management and infection control are critical. On February 4, 2020, the Secretary of Health and Human Services (HHS) confirmed a public health emergency (PHE) and that circumstances exist justifying the authorization of emergency use of in vitro diagnostics for detection of SARS-CoV-2 [2],[3]. Rapid detection of COVID-19 cases requires wide availability of diagnostic testing to control the emergence of this rapidly spreading, severe illness.

The SARS-CoV-2 genome encodes approximately 25 proteins that are required for infection and replication, including four major structural proteins spike (S), envelope (E), membrane (M), and nucleocapsid (N) protein [4],[5]. ELISA or CLIA assays are the most frequently used format for serological testing with an average turnaround time of 2 to 5 hours. A rapid diagnostic test (RDT) is a simple test based on lateral flow immunoassay (LFIA) technology and can potentially be administered as a point-of-care (POC) test or self-test. RDT test strips can provide reliable qualitative detection and differentiation of IgM, IgG and/or IgA antibodies against SARS-CoV-2 from a drop of blood. It is important to note that donor antibodies (IgG, IgM, or IgA) are produced against a specific SARS-CoV-2. In case of an RDT such as lateral flow immunoassays (LFIA), when a specimen followed by assay buffer is added to the sample well, and IgM/IgG/IgA antibodies are present, these antibodies bind to colloidal gold conjugated to recombinant COVID-19 antigens (e.g. SARS-CoV-2 spike or nucleocapsid antigen) immobilized on a nitrocellulose strip. The antibody/COVID-19 conjugate complex migrates through a nitrocellulose membrane by capillary action to a line of the corresponding immobilized antibody (anti-human IgM, anti-human IgG or anti-human IgA) where it is trapped forming a colored band which confirms a reactive test result. Absence of a colored band in the test region indicates a non-reactive test result [6].

During the COVID-19 pandemic, the majority of emerging RDTs receiving Emergency Use Authorization (EUA) approval by the Food and Drug Administration (FDA) for qualitative detection and differentiation of IgM and IgG antibodies to SARS-CoV-2 were only approved for use in human venous whole blood, plasma or serum by laboratories certified under the Clinical Laboratory Improvement Amendments of 1988 (CLIA). As of May 29, 2020, the FDA provided EUA to a COVID-19 IgG/IgM rapid test cassette serology test manufactured by Healgen® for use in human venous whole blood, plasma from anticoagulated blood (Li+ heparin, K2EDTA and sodium citrate), or serum [7]–[9]. As of June 19, 2020, the FDA provided EUA to a similar product, the LYHER® Novel Coronavirus (2019-nCoV) IgM/IgG Antibody Combo Test Kit (Colloidal Gold) for use in human serum and plasma (Li+ heparin, K 2EDTA or sodium-citrate) [10].

In this study we summarize performance characteristics of the Healgen® COVID-19 IgG/IgM lateral flow immunoassay rapid cassette to the LYHER® Novel Coronavirus (2019-nCoV) IgM/IgG Antibody Combo Test Kit, both LFIAs specifically targeting antibodies binding the SARS-CoV-2 spike S1 antigen. In order to determine performance of a different specimen source, simultaneous application of whole blood finger-stick specimens (n = 32) was performed over 5 different days, with daily quality controls consisting of serum previously verified to be positive or negative by COVID-19 IgG/IgM ELISA testing.

## 2 Materials and Methods

### 2.1 Study population

This study was approved by the Alcala Pharmaceutical Inc. Institutional Review Board (IORG0010127) in consideration of the Code of Ethics of the World Medical Association (Declaration of Helsinki). Specimen collections were performed at the phlebotomy collection site at Alcala Testing and Analysis Services, 3703 Camino del Rio South, San Diego, CA, 92108 by certified phlebotomists. Whole blood specimen rapid cassette testing was conducted from 08/14/2020 to 08/26/2020 at Alcala Testing and Analysis Services. All subjects were recruited by phone or e-mail at the San Diego Comprehensive Pain Management Clinic, 3703 Camino del Rio South, San Diego, CA, 92108. Healthcare workers from the clinic or volunteers in any setting (symptomatic or asymptomatic) could enroll without a prior COVID-19 test. All specimens derived from human subjects were de-identified of their health information as defined by Health Insurance Portability and Accountability Act (HIPAA) guidelines. Thirty-two volunteers took part in a whole blood finger-stick collection with simultaneous application of the specimen to the Healgen® or LYHER® COVID-19 IgG and IgM antibody rapid cassette.

### 2.2 Materials, collection and test kit components

Whole blood finger-sticks for Healgen® and LYHER® lateral flow rapid cassette testing were collected with kits containing 21G (2.2. mm depth) high flow safety lancets (One-Care®, Irvine, CA), alcohol prep pad, gauze and band-aids. Use of digital timers ensured rapid cassette image interpretation at the recommended 10-minute mark. Deidentified, positive and negative control serum for IgG/IgM COVID-19 antibodies was made available for daily quality control (QC) testing of both rapid cassettes. QC serum was verified by the EDI™ Novel Coronavirus COVID-19 IgG/IgM ELISA diagnostic kit as described previously in Baig et al. [11].

### 2.3 Sample processing and storage

Healgen® and LYHER® lateral flow rapid cassette kits for COVID-19 IgG and IgM antibody detection were used for testing individuals by whole blood finger-stick at the Alcala Labs collection site. 3 donors who tested for positive for IgG and/or IgM COVID-19 antibodies also provided nasopharyngeal swabs for confirmation COVID-19 by RT-PCR (Nucleic acid amplification technique or NAAT). RT-PCR analysis was performed by Luminex MAGPIX NxTAG® CoV Extended Panel as previously described [11].

### 2.4 Healgen® lateral flow rapid cassette kit procedure and result interpretation

Sample buffer was equilibrated at room temperature (15-30°C) prior to testing. Rapid cassettes and the 5 μL mini plastic dropper were removed from the sealed foil pouch and placed on a clean and level surface. After cleansing the finger stick collection site with the alcohol prep pad, whole blood was drawn using the 21G lancet and whole blood specimen (or serum quality control material) applied into the sample well (S) of the rapid cassette with the mini plastic dropper. Two drops (approximately 80 μL) of sample buffer were added to buffer well (B) while avoiding air bubbles (**Fig 1a**). An additional drop of the sample buffer was added within 2 minutes if the red color did not move across the test window. The result was read in ten minutes. Result interpretation after fifteen minutes was avoided.

**Fig 1.**
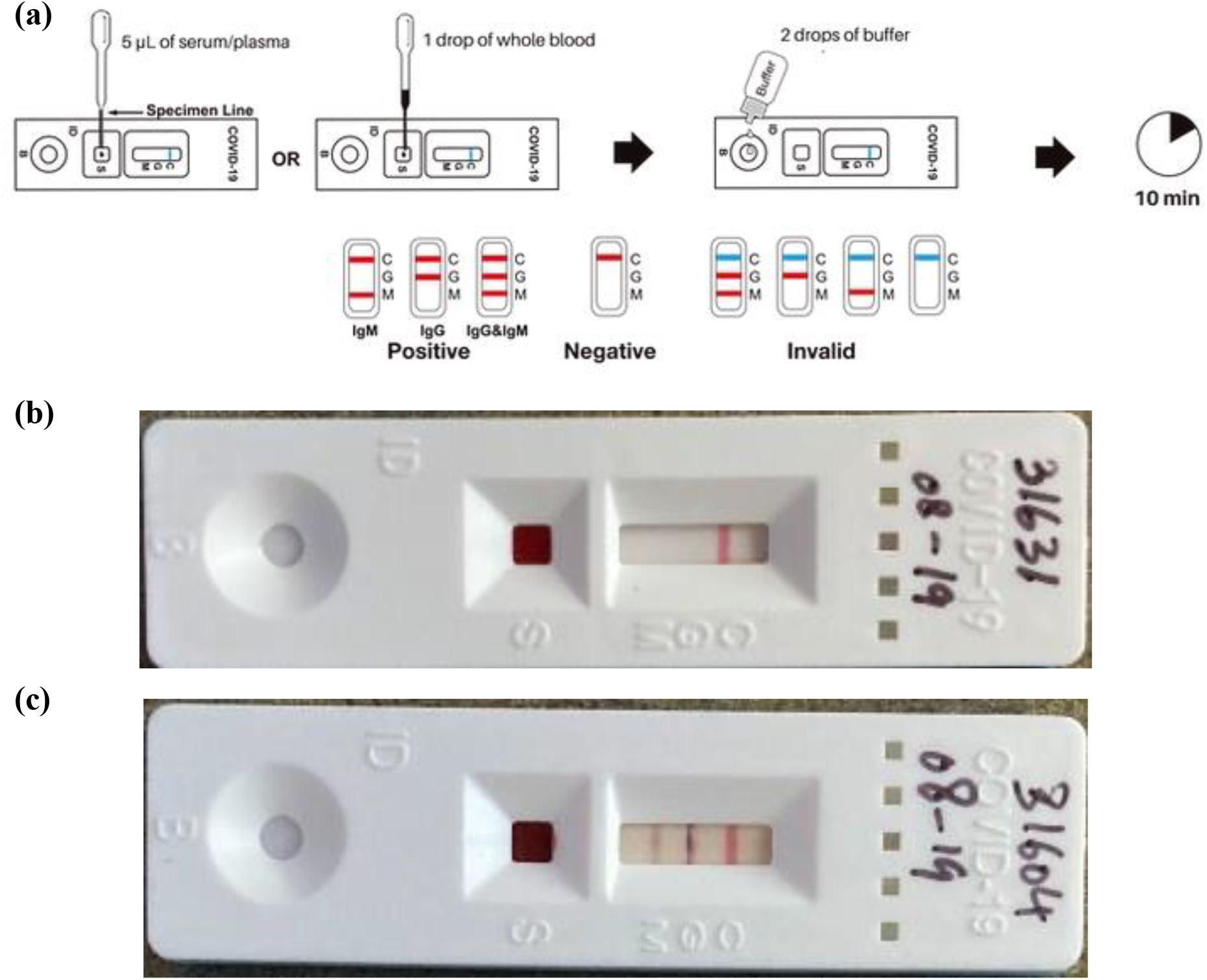
(a) Healgen® lateral flow rapid cassette kit procedure and result interpretation. (b) Negative result Healgen® rapid cassette. (c) IgG and IgM positive result Healgen® rapid cassette.

If the internal control in the control line region (C) changed from blue to red and no line appeared in the test line regions M or G results were interpreted as negative (**Fig 1b**). IgM positive was determined if the colored line in the control line region (C) changed from blue to red, and a colored line appeared in region M, indicating the presence of IgM anti-SARS-CoV-2 antibodies. The test was considered IgG positive, if the colored line in the control line region (C) changed from blue to red, and a colored line appeared in test line region G. This result indicates the presence of IgG anti-SARS-CoV-2 antibodies. For both IgG and IgM to be considered positive two colored lines were visible in test line regions M and G in combination with the control line region (C) changing from blue to red. The test results indicate the presence of both IgM and IgG anti-SARS-CoV-2 antibodies (**Fig 1c**). An invalid result was not observed, in this case the control line may be partially red or completely fail to change to red. Insufficient specimen volume, incorrectly stored or expired rapid cassette lots are the most likely reasons for control line failure.

### 2.5 LYHER® lateral flow rapid cassette kit procedure and result interpretation

Sample buffer was equilibrated at room temperature (15-30°C) prior to testing. Rapid cassettes (no mini plastic dropper provided) were removed from the sealed foil pouch and placed on a clean and level surface. Upon cleansing the finger stick collection site with the alcohol prep pad, whole blood was drawn using the 21G lancet, whole blood specimen (or serum quality control material) was applied to the sample well (S) of the rapid cassette using a 200 μL transfer pipette. Two drops (approximately 80 μL) of sample buffer was added to the (S) well while avoiding air bubbles. The LYHER® lateral flow rapid cassette features no separate buffer well (**Fig 2a**). The result was read in ten minutes. Result interpretation after fifteen minutes was avoided.

**Fig 2.**
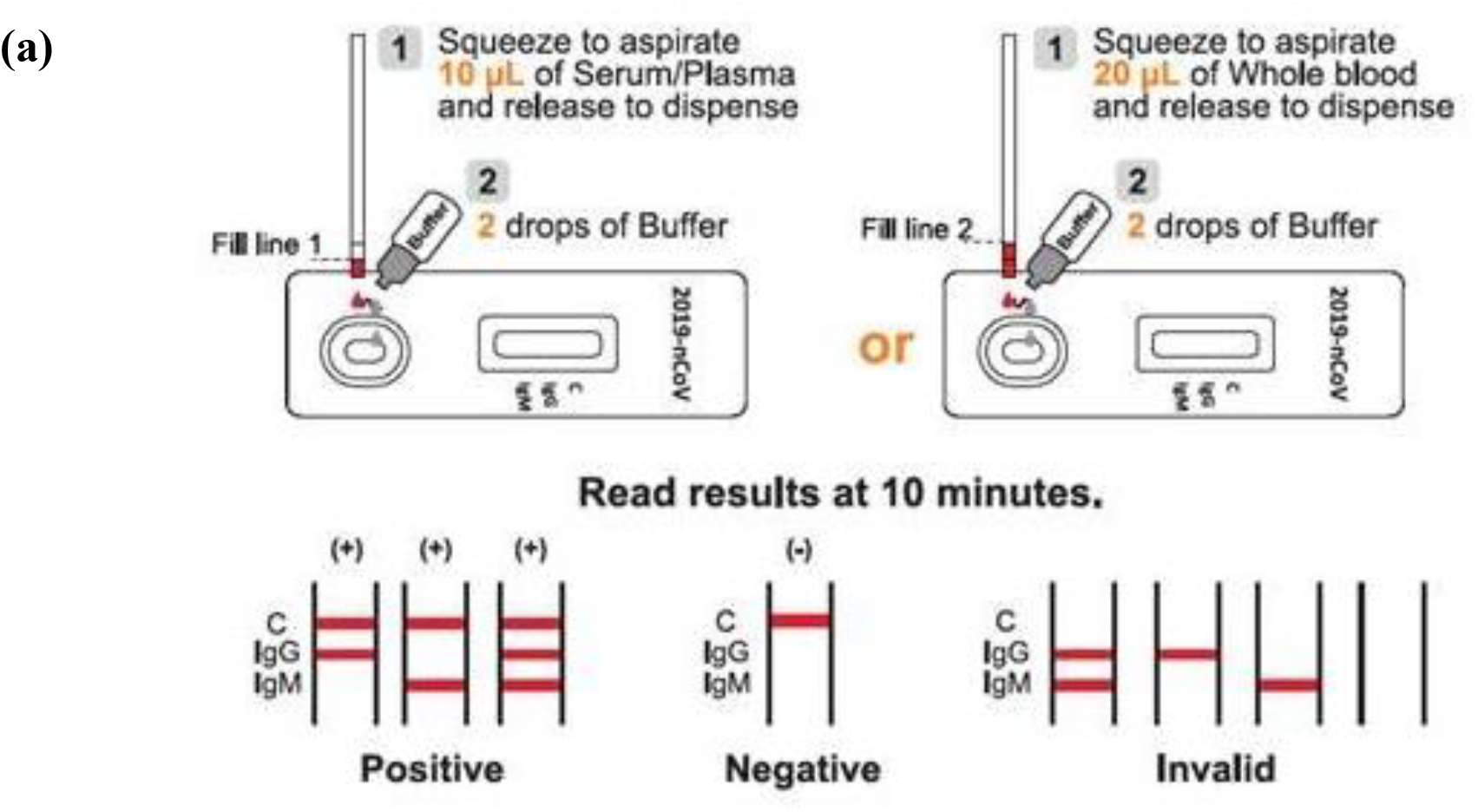
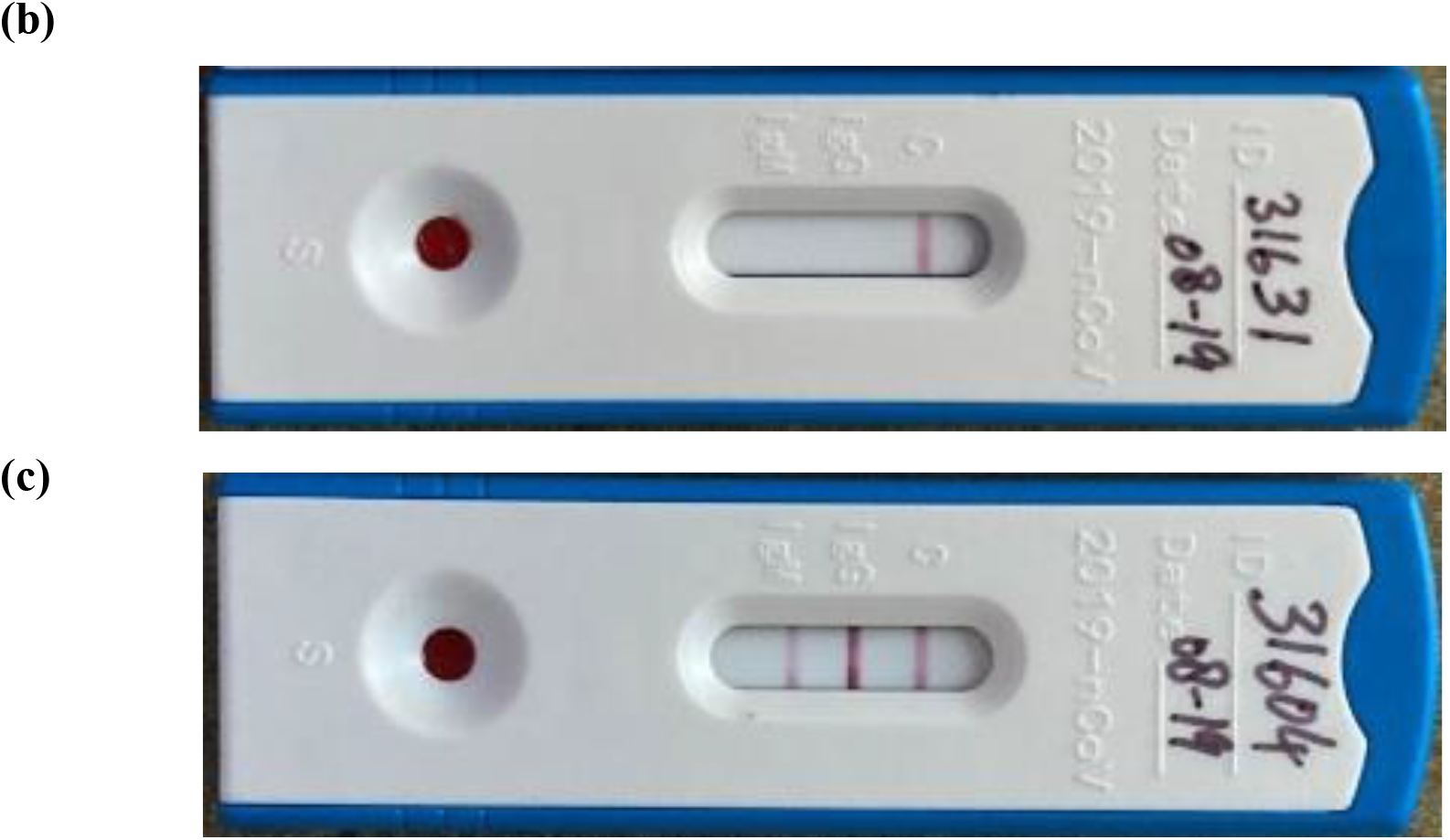
(a) LYHER® lateral flow rapid cassette kit procedure and result interpretation. (b) Negative result LYHER® rapid cassette. (c) IgG and IgM positive result LYHER® rapid cassette.

If the internal control line on the nitrocellulose membrane (control line region (C)) changed from white to red and no line appeared in the test line regions M or G results were interpreted as negative (**Fig 2b**). IgM positive was determined if the colored line in the control line region (C) changed from white to red, and a colored line appeared in region M, indicating the presence of IgM anti-SARS-CoV-2 antibodies. The test was considered IgG positive, if the colored line in the control line region (C) changed from white to red, and a colored line appeared in test line region G. This result indicates the presence of IgG anti-SARS-CoV-2 antibodies. For both IgG and IgM to be considered positive two colored lines were visible in test line regions M and G in combination with the control line region (C) changing from white to red. The test results indicate the presence of both IgM and IgG anti-SARS-CoV-2 antibodies (**Fig 2c**). An invalid result was not observed, in this case the control line may be partially red or completely fail to change to red. Insufficient specimen volume, incorrectly stored or expired rapid cassette lots are the most likely reasons for control line failure.

### 2.6 Quality Controls

An internal control band is included in both the Healgen® and LYHER® lateral flow immunoassays. A red line appears in the control region (C) if sufficient specimen volume was applied, and the colorimetric lateral flow immunoassay is performing as expected. It is recommended by the FDA that positive and negative controls be tested as a good laboratory practice to confirm the test procedure and to verify proper test performance for both EUA-approved products described here. In this study pretested IgG or IgG, IgM positive serum control material and pretested IgG, IgM negative serum control material was pipetted onto 2 separate cassettes for every Healgen® and LYHER® lateral flow rapid cassette batch tested within 24 hours.

## 3. Results and Discussion

The Healgen® COVID-19 IgG/IgM Rapid Test Cassette was originally evaluated in China by testing a total of 191 plasma (K2EDTA) clinical samples and a study by the Frederick National Laboratory for Cancer Research (FNLCR) using frozen serum and plasma specimens, with 30 positives confirmed by RT-PCR and 80 negative specimens derived from collections before 2020 (n = 110). Alcala Labs performed a similar study on 75 clinical specimens comparing whole blood finger-stick collections to a simultaneous venipuncture draw serum collection measured by IgG/IgM ELISA [11] (**Table 1**). All 3 studies showed that a whole blood finger-stick collection versus venipuncture serum draws performs equally well in regard to combined sensitivity (100%) and 1% higher in combined specificity. Overall, the combined positive predictive value (PPV) at a presumed prevalence of 5% increased by 10% and the combined negative predictive value (PPV) at a presumed prevalence of 5% was maintained at 100% (**Table 1**): Clinical performance of the LYHER® Novel Coronavirus (2019-nCoV) IgM/IgG Antibody Combo Test Kit was initially established from 527 specimens collected at four medical centers in Hubei Province, China, from patients who tested positive by an RT-PCR test or patients who recovered from COVID-19, and from individuals who had contact with COVID-19 patients but tested negative by RT-PCR testing. Among the 178 specimens that were PCR positive, 90 were from patients who were still in quarantine and 88 were from recovered patients. Among the 349 negative samples, 239 originated from patients with coronavirus infections not caused by SARS-CoV-2, and 110 patients with other respiratory tract infections [12] (**Table 2**).

**Table 1:** Summary Statistics comparison for the Healgen® COVID-19 IgG/IgM Rapid Test Cassette between the China study, FNLCR study and previous whole-blood finger stick study (Baig et al.):

|  | Summary Statistics |  |  |  |
| --- | --- | --- | --- | --- |
|  | Combined Sensitivity/PPA | Combined Specificity/NPA | Combined PPV for prevalence = 5% | Combined NPV for prevalence = 5% |
| China study (n = 191) | 96.7% (87/90) | 97% (98/101) | 62.9% | 99.8% |
| FNLCR study (n = 110) | 100% (30/30) | 97.5% (78/80) | 67.8% | 100% |
| Baig et al. (n = 75) | 100% (11/11) | 98.5% (63/64) | 77.8% | 100% |

**Table 2:** Summary Results and Statistics for the LYHER® Novel Coronavirus (2019-nCov) IgM/IgG Antibody Combo Test Kit validated by testing plasma (K2EDTA) clinical samples from individual patients exhibiting symptoms or confirmed negative by RT-PCR at four sites in China (n = 527):

| Method | Comparator Method: RT-PCR |  |  |  |
| --- | --- | --- | --- | --- |
| LYHER®<br>Antibody<br>IgG/IgM<br>Combo Test |  | Positive | Negative | Total |
| Positive | IgG+/IgM+ | 144 | 3 | 147 |
|  | IgG-/IgM+ | 30 | 0 | 30 |
|  | IgG+/IgM- | 1 | 0 | 1 |
| Negative | IgG-/IgM- | 0 | 347 | 347 |
| **Subtotal** |  | 178 | 349 | 527 |
| Summary Statistics |  |  |  |  |
| Measure | Estimate |  | 95% Confidence Interval |  |
| IgG Sensitivity/PPA | 81.4% (145/178) |  | (75.0%; 86.9%) |  |
| IgG Specificity/NPA | 99.4% (347/349) |  | (97.9%; 99.9%) |  |
| IgM Sensitivity/PPA | 97.7% (174/178) |  | (94.4%; 99.4%) |  |
| IgM Specificity/NPA | 99.4% (347/349) |  | (97.9%; 99.9%) |  |
| Combined Sensitivity/PPA | 97.7% (174/178) |  | (94.4%; 99.4%) |  |
| Combined Specificity/NPA | 99.4% (347/349) |  | (97.9%; 99.9%) |  |
| Combined PPV for prevalence = 5% | 89.6% |  |  |  |
| Combined NPV for prevalence = 5% | 99.9% |  |  |  |

The LYHER Novel Coronavirus (2019-nCoV) IgM/IgG Antibody Combo Test Kit (Colloidal Gold) was also tested on 6/10/2020 at the Frederick National Laboratory for Cancer Research (FNLCR) sponsored by the National Cancer Institute (NCI). The test was validated against a panel of previously frozen samples consisting of 30 SARS-CoV-2 antibody-positive serum samples and 80 antibody-negative serum and plasma samples. Each of the 30 antibody -positive samples were confirmed with a nucleic acid amplification test (NAAT) and both IgM and IgG antibodies were confirmed to be present in all 30 samples [13] (**Table 3**):

**Table 3:**
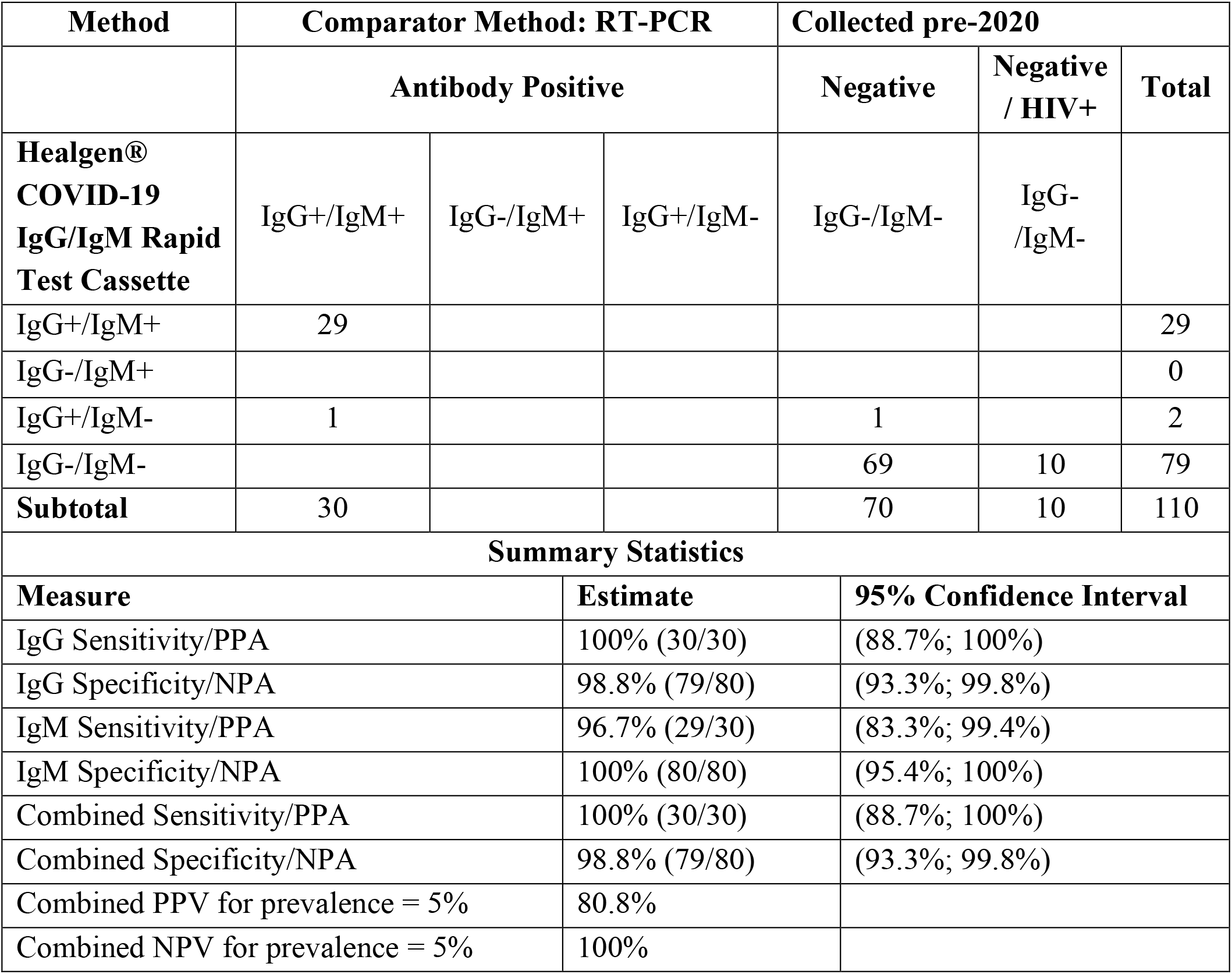
Summary Results and Statistics for the LYHER® COVID-19 IgG/IgM Rapid Test Cassette validated at the Frederick National Laboratory for Cancer Research (FNLCR) from frozen venipuncture serum and plasma samples (n = 110):

The Healgen® COVID-19 IgG/IgM Rapid Test Cassette was also originally evaluated using by the Frederick National Laboratory for Cancer Research (n = 110 serum or plasma specimens) with positives confirmed by RT-PCR, a direct comparison between the 2 studies at the FNLCR in **Table 4** shows equal performance between the 2 rapid cassettes for serum or plasma specimens in regard to combined sensitivity, and 1% higher combined specificity for the LYHER® LFIA product.

**Table 4:** Summary Statistics comparison for the Healgen® and LYHER® COVID-19 IgG/IgM Rapid Test Cassette at the FNLCR:

|  | Summary Statistics |  |  |  |
| --- | --- | --- | --- | --- |
|  | Combined Sensitivity/PPA | Combined Specificity/NPA | Combined PPV for prevalence = 5% | Combined NPV for prevalence = 5% |
| Healgen® study (n = 110) | 100% (30/30) | 97.5% (78/80) | 67.8% | 100% |
| LYHER® study (n = 110) | 100% (30/30) | 98.8% (79/80) | 80.8% | 100% |

To evaluate whole blood derived from finger-sticks more applicable to remote, point-of-care settings, Alcala Labs compared 32 whole blood specimens each applied simultaneously to both the Healgen® and LYHER® COVID-19 IgG/IgM rapid test cassette over 5 different days with daily positive and negative controls applied to each of the cassettes (**Table 5**). Daily quality controls consisted of serum verified to be positive or negative by the EDI™ Novel Coronavirus COVID-19 IgG/IgM ELISA validated at Alcala Labs.

**Table 5:**
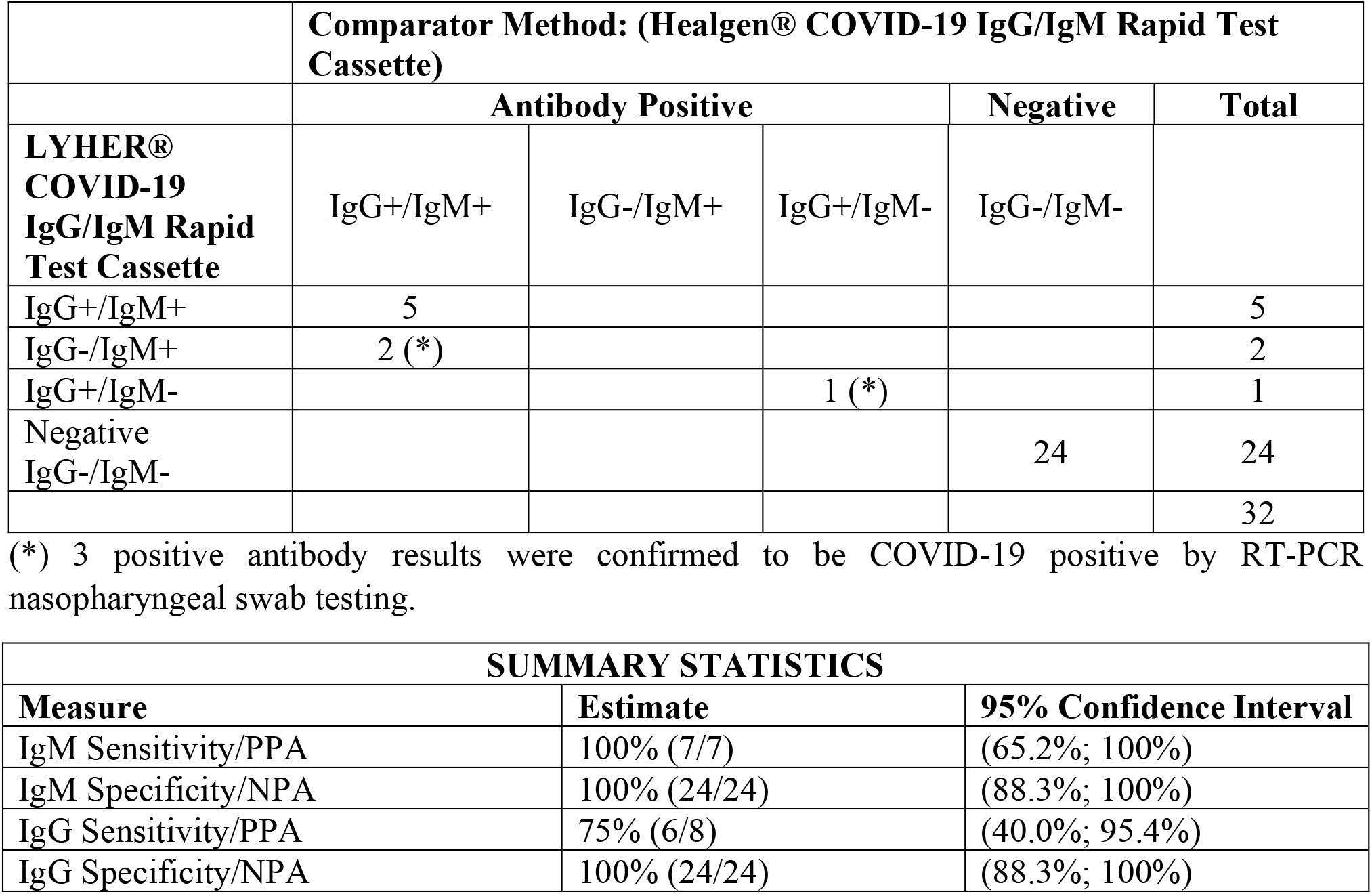
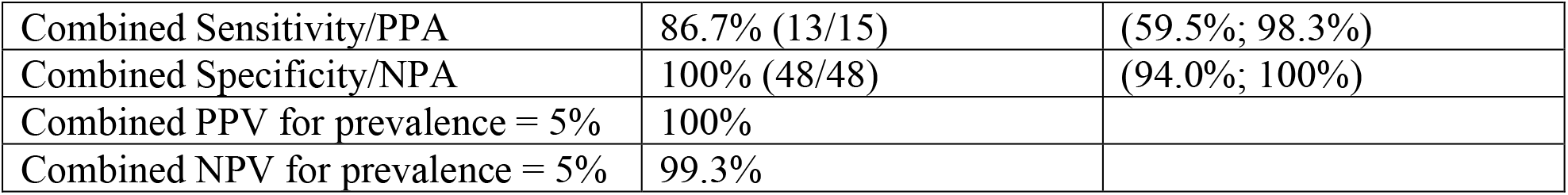
Summary Results and Statistics comparing the Healgen® versus LYHER® COVID-19 IgG/IgM Rapid Test Cassette for the whole blood finger-stick sample type (n = 32):

|  | Comparator Method: (Healgen® COVID-19 IgG/IgM Rapid Test Cassette) |  |  |  |  |
| --- | --- | --- | --- | --- | --- |
|  | Antibody Positive |  |  | Negative | Total |
| LYHER® COVID-19 IgG/IgM Rapid Test Cassette | IgG+/IgM+ | IgG-/IgM+ | IgG+/IgM- | IgG-/IgM- |  |
| IgG+/IgM+ | 5 |  |  |  | 5 |
| IgG-/IgM+ | 2 (*) |  |  |  | 2 |
| IgG+/IgM- |  |  | 1 (*) |  | 1 |
| Negative IgG-/IgM- |  |  |  | 24 | 24 |
|  |  |  |  |  | 32 |

(\*) 3 positive antibody results were confirmed to be COVID-19 positive by RT-PCR nasopharyngeal swab testing.
| SUMMARY STATISTICS |  |  |
| --- | --- | --- |
| Measure | Estimate | 95% Confidence Interval |
| IgM Sensitivity/PPA | 100% (7/7) | (65.2%; 100%) |
| IgM Specificity/NPA | 100% (24/24) | (88.3%; 100%) |
| IgG Sensitivity/PPA | 75% (6/8) | (40.0%; 95.4%) |
| IgG Specificity/NPA | 100% (24/24) | (88.3%; 100%) |
| Combined Sensitivity/PPA | 86.7% (13/15) | (59.5%; 98.3%) |
| Combined Specificity/NPA | 100% (48/48) | (94.0%; 100%) |
| Combined PPV for prevalence = 5% | 100% |  |
| Combined NPV for prevalence = 5% | 99.3% |  |

Based on this data the LYHER® rapid cassette shows a combined sensitivity of 86.7% and combined specificity of 100% in whole blood finger-stick specimens versus the Healgen® rapid cassette. The negative predictive value (NPV) at 5% prevalence was determined at 99.3% and the positive predictive value (PPV) at 100%.

## 4. Conclusion

This study confirmed that utilizing the LYHER® COVID-19 IgG/IgM rapid cassette by applying whole blood finger-stick derived specimens instead of serum or plasma from a venipuncture draw shows a combined sensitivity of 86.7% and combined specificity of 100% versus the Healgen® COVID-19 IgG/IgM rapid cassette. Clearly, performance of both COVID-19 IgG/IgM rapid diagnostic tests using LFIA technology is highly sensitive and specific in venipuncture serum or plasma (both at 99.4-100% sensitivity and 97.5-98.8% specificity). As has been shown in the Baig et al. study (n = 75) [11], Healgen® LFIA technology is 100% sensitive in whole blood finger-stick versus serum. LYHER® LFIA technology is 13.3% less sensitive in whole blood finger-stick but maintains 100% specificity. This study may skew sensitivity to a lower percentage due to a relatively smaller sample size of positive whole blood specimens (n = 8) and overall low sample size (n = 32). However, the negative predictive value (NPV) at 5% prevalence was determined at 99.3% and the positive predictive value (PPV) at 100% showing both rapid diagnostic tests can be reliably applied in remote settings on whole blood finger-stick specimens.

## Data Availability

All data related to this study is contained within the manuscript and all volunteer data has been deidentified as per HIPAA guidelines.

## Author Contributions

Conceptualization, C.T. and D.J.S; methodology, C.T., I.B.; validation, C.T., M.J.C.M; formal analysis, R.S.Z., M.J.C.M., C.T.; Resources, D.J.S.; writing - original draft preparation, C.T., writing - review and editing, C.T., M.J.C.M; supervision, D.J.S.; project administration, C.T.

## Acknowledgments

The authors would like to thank the San Diego Comprehensive Pain Management Clinic (SDCPMC) and associated staff for donor collections performed during this study.

